# A network-informed analysis of SARS-CoV-2 and hemophagocytic lymphohistiocytosis genes’ interactions points to Neutrophil Extracellular Traps as mediators of thrombosis in COVID-19

**DOI:** 10.1101/2020.07.01.20144121

**Authors:** Jun Ding, David Earl Hostallero, Mohamed Reda El Khili, Gregory Fonseca, Simon Millette, Nuzha Noorah, Myriam Guay-Belzile, Jonathan Spicer, Noriko Daneshtalab, Martin Sirois, Karine Tremblay, Amin Emad, Simon Rousseau

## Abstract

Abnormal coagulation and an increased risk of thrombosis are features of severe COVID-19, with parallels proposed with hemophagocytic lymphohistiocytosis (HLH), a life-threating condition associated with hyperinflammation. The presence of HLH was described in severely ill patients during the H1N1 influenza epidemic, presenting with pulmonary vascular thrombosis. We tested the hypothesis that genes causing primary HLH regulate pathways linking pulmonary thromboembolism to the presence of SARS-CoV-2 using novel network-informed computational algorithms. This approach led to the identification of Neutrophils Extracellular Traps (NETs) as plausible mediators of vascular thrombosis in severe COVID-19 in children and adults. Taken together, the network-informed analysis led us to propose the following model: the release of NETs in response to inflammatory signals acting in concert with SARS-CoV-2 damage the endothelium and direct platelet-activation promoting abnormal coagulation leading to serious complications of COVID-19. The underlying hypothesis is that genetic and/or environmental conditions that favor the release of NETs may predispose individuals to thrombotic complications of COVID-19 due to an increase risk of abnormal coagulation. This would be a common pathogenic mechanism in conditions including autoimmune/infectious diseases, hematologic and metabolic disorders.

## Introduction

Early reports from China^1,2^ and France^3,4^, have highlighted abnormal coagulation and a high risk of thrombosis in severe COVID-19. Evidence of both micro-thrombosis^5^ and macro-thrombosis^6^ have emerged. Understanding the mechanism by which SARS-CoV-2 infections lead to coagulopathy is an important goal of the current research effort to properly identify and treat individuals at risk of severe complications of COVID-19.

A strong pro-inflammatory but ineffective anti-viral response contributes to COVID-19^7^. This can lead to a hyperinflammatory state described in a subset of COVID-19 adult patients with severe and often deadly complications^8^. This hyperinflammation in severe COVID-19 patients includes elevated levels of C-reactive protein (CRP), ferritin, fibrinogen and D-dimers^9–11^. Parallels have been drawn between hematologic cytokine storms and severe COVID-19 pointing to hemophagocytic lymphohistiocytosis (HLH) as a putative link^12^. HLH is a life-threating condition associated with hyperinflammation resulting in varying clinical manifestations such as fever, organomegaly, coagulopathy, cytopenia, neurologic dysfunction, and elevations of acute phase reactants. The presence of HLH was also described in severely ill patients during the novel H1N1 influenza epidemic of 2009, presenting with peripheral pulmonary vascular thrombosis^13– 16^. Interestingly, the post-mortem lungs from patients with COVID-19 showed severe endothelial injury associated with the presence of intracellular virus and alveolar capillary microthrombi. This was 9 times more prevalent in COVID-19 than H1N1 lungs analysed following autopsy^17^. Accordingly, in one study, COVID-19 Acute Respiratory Distress Syndrome (ARDS) developed significantly more thrombotic complications than non-COVID-19 ARDS^4^. HLH has also been observed in children suffering from Kawasaki Disease (KD), a form of systemic vasculitis in children^18^ A 30-fold increase of incidence of KD-like illness has been observed in Bergamo, Italy following the SARS-CoV-2 outbreak^19^, as well in Paris, France, preprint^20^ and the UK^21^. In the KD-like illness described in the Italian study, half of the patients were diagnosed with Macrophage-Activation Syndrome (MAS), a condition closely related to HLH. Interestingly, hyperinflammation, rather than the hemophagocytosis appears to be the driving cause of pathology^22^. Therefore, vascular injury resulting from hyperinflammation in the alveoli in response to SARS-CoV-2 may be an important pathophysiological mechanism of severe illness in children and adults.

Based on the above reports, we hypothesized that genes causing the genetic forms of HLH and associated syndromes regulate pathways linking the risk of pulmonary microvascular thromboembolism to the presence of SARS-CoV-2. To test this hypothesis quickly in view of the world-wide pandemic, we used network-informed approaches, including a novel computational tool to explore the link between HLH genes and a network of human proteins identified to interact with SARS-CoV-2^23^. This approach led to the identification of Neutrophils Extracellular Traps (NETs) as plausible mediators of vascular thrombosis in severe COVID-19.

## Results

### HLH genes are significantly enriched within the SARS-CoV-2 host protein interactome

In the case of the SARS-Cov-2 pandemic, with widespread impact across the world, there is an urgency that requires the adaptation of different strategies to understand COVID-19. In this paper, we exploited the knowledge existing within protein interaction networks to identify the molecular pathways underpinning thrombotic complications of COVID-19 using advanced computational algorithms. As described in the introduction, a subset of patients suffering from severe complications of COVID-19 present clinically with symptoms similar to HLH. Therefore, we have assembled a list of candidate genes responsible for primary HLH and associated syndromes to explore their relationships with COVID-19^24,25^ (**Supplementary Table S1**).

The first question asked was whether these HLH genes had potential interactions with SARS-CoV-2. We assembled a protein interaction network between the SARS-CoV-2 host interaction protein network recently published^23^ and the HLH genes using an algorithm that we created for this purpose, GeneList2COVID19. The algorithm establishes the shortest path between the candidate genes and the known host interacting proteins with SARS-CoV-2 and calculates an overall connectivity score for the network (a smaller value represents a greater connectivity) (**Fig 1 and Supplementary Table S1)**. We computationally validated the predictions of the GeneList2COVID19 to identify significant interactions. To demonstrate that the method can assign significant connectivity scores to genes associated with COVID-19, we obtained a list of 10 confirmed COVID-19 related genes^26^, which are differentially expressed in severe COVID19 patients (**Supplementary Table S1**). We then calculated the “COVID-19” connectivity score for those 10 genes (SA) as well as all the genes (SB) using GeneList2COVID19. We found that SA is significantly (p-value=0.017) smaller than SB, which indicates that those 10 COVID-19 related genes are indeed “significantly connected” to SARS-CoV-2 proteins (**Fig. 2**). To show the specificity of the method, we also calculated the “COVID19” connectivity score for 100 randomly selected genes (SC) and compared it to the connectivity score of all genes (SB). We found that SC is NOT significantly smaller than B (the background) (p-value=0.106) (**Fig. 2**). In other words, those 100 random genes are not “significantly connected” to SARS-CoV2 proteins, which reflects the fact that those genes were randomly picked. As an additional control, we repeated the analysis using genes linked to male infertility^27^, a condition that has not been associated with COVID-19 (**Supplementary Table S1**). The connectivity score was not significantly different from all other genes (p-value=0.872), further demonstrating the specificity of the GeneList2COVID19, which is not restricted to random genes but can also discriminate gene lists associated with other conditions (**Fig. 2**). After the method was validated, we compared the “connectivity” score for HLH genes listed above with all genes that connect to SARS-CoV-2 proteins through our assembled protein-protein interaction network (**Fig. 2**). We found that the score for the HLH marker genes is significantly smaller compared to all other genes (p-value=0.0082, one-sided rank sum test) (**Fig. 2**). As an additional control, we compared the HLH genes to a list of vascular angiogenesis genes linked to both H1N1 and SARS-CoV-2 pulmonary infections^17^ (**Supplementary Table S1 & Fig. 2**). The HLH genes’ connectivity score was smaller, which means that those genes had closer theoretical interactions to SARS-CoV-2. This suggests that HLH genes and their associated pathways are of high interest in the study of SARS-CoV-2 infections.

**Figure 1.**
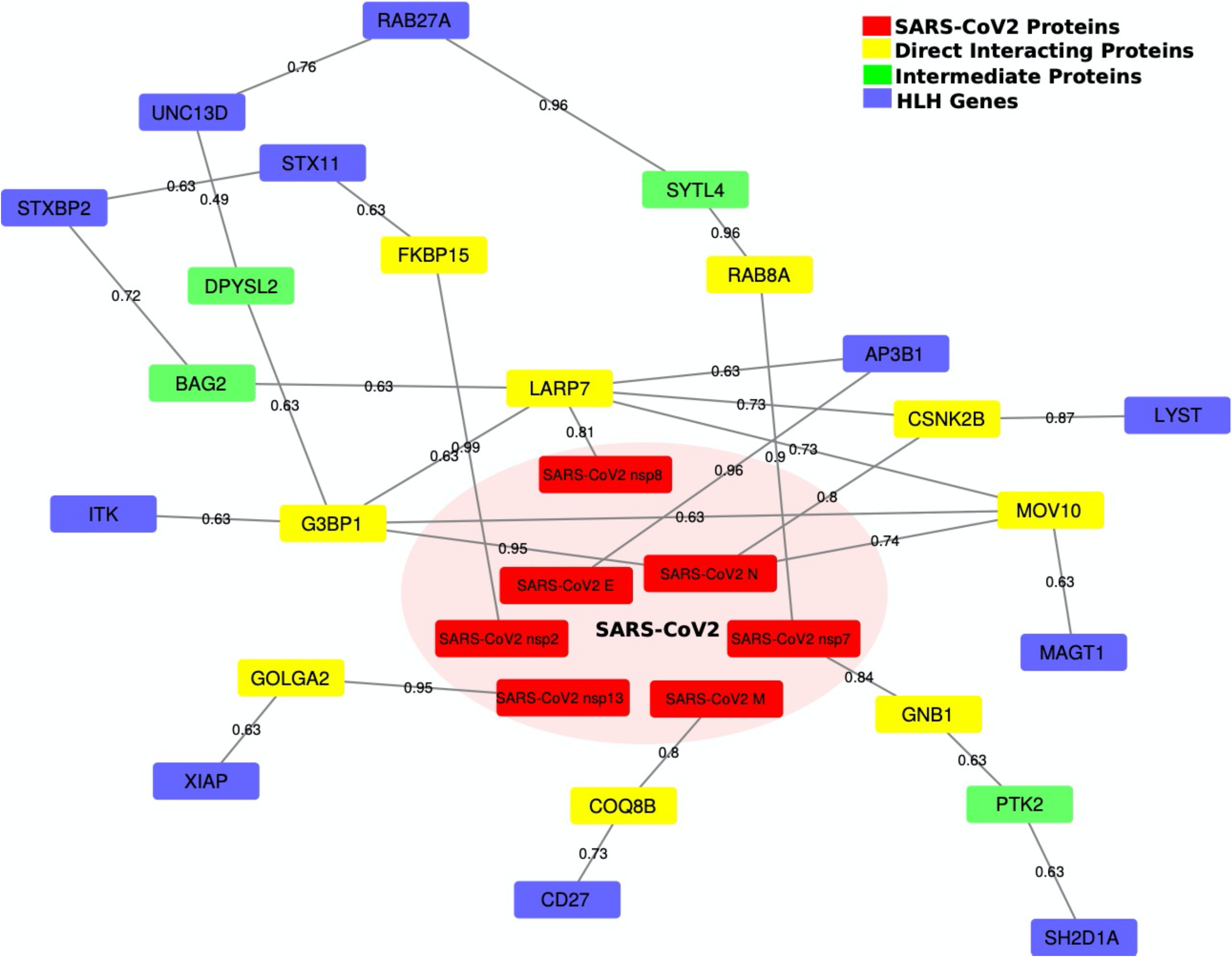
Reconstructed network paths from SARS-CoV-2 proteins to HLH genes. This network shows all the paths connecting the SARS-CoV-2 proteins to the HLH proteins (genes). The red nodes represent the SARS-CoV-2 proteins, the yellow nodes are the human host proteins that directly interact with SARS-CoV-2 proteins, the green nodes are the intermediate interacting host proteins, and the blue nodes denote the target HLH proteins (genes). The edge weights in the network represent the interaction strength (or probability).

**Figure 2.**
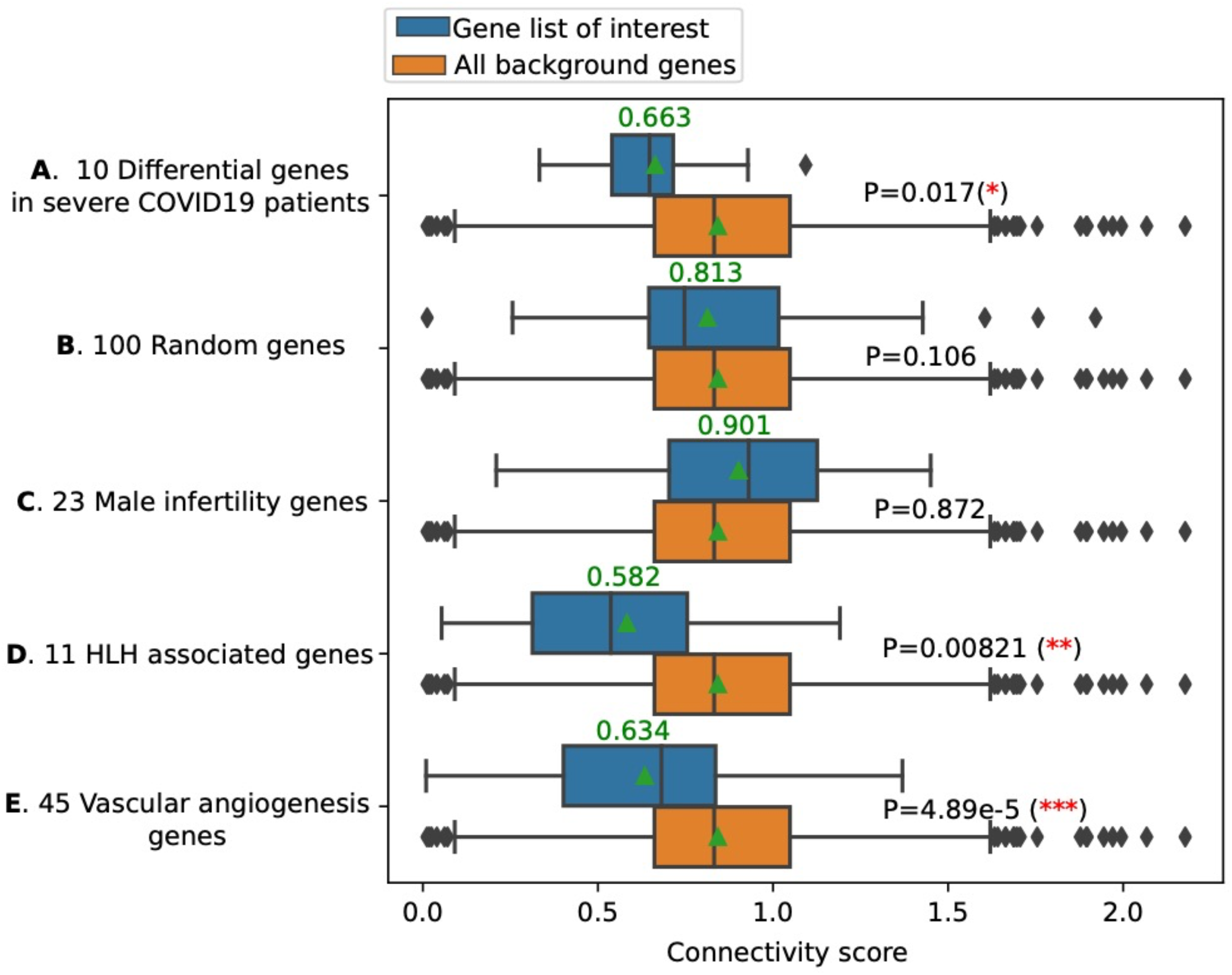
HLH genes are significantly enriched within the SARS-CoV-2 host protein interactome. A connectivity score was calculated for each of the genes of interest (e.g. HLH genes in this work). We further analyzed the network connectivity of all genes to the SARS-CoV2 proteins (or randomly picked genes). With these two analyses, we ended up with two lists of connectivity scores: *SA* (for HLH genes) and *SB* (for all background genes). Then, we calculated the statistical significance (p-value) using a one-sided Mann-Whitney rank test to determine whether *SA* is significantly smaller than *SB* (stronger connectivity). *SA* significant p-value implies that the list of proteins (genes) of interest is “significantly connected” to the SARS-CoV2 proteins. **A**) A list of 10 known COVID19 related genes (differential genes in severe COVID19 patients) have statistically stronger connections to the SARS-CoV-2 proteins compared with all background genes (p-value=0.017). **B)** A list of 100 random genes does not “significantly” connect to the SARS-CoV-2 proteins. **C)** The 23 Male infertility genes do not “significantly” connect to the SARS-CoV-2 proteins. **D)** The 11 HLH genes have statistically (p-value=0.00821) stronger connections to the SARS-CoV-2 proteins (compared with all background genes). **E)** A list of 45 vascular angiogenesis genes linked to both H1N1 and SARS-CoV-2 pulmonary infections significantly (p-value=4.89e-5) connect to the SARS-CoV-2 proteins. The 11 HLH genes have the smallest mean/median connectivity score compared to all the gene lists analyzed. Please note that the p-values here only indicate whether the input gene lists have significantly smaller connectivity scores than all the background genes, and they could be affected by the size of the gene list. To compare the strength of the “connectivity” of input gene lists to SARS-CoV-2 proteins, we should also look at the mean (represented by a green triangle) and the median (represent by a vertical line) connectivity scores.

### Differential expression of HLH genes in health conditions related to COVID-19

We next investigated whether the expression of HLH genes in lung tissue were highly regulated (up or down) in conditions associated with COVID-19 (Sex, Smoking, Lung Cancer, Chronic Obstructive Pulmonary Disease (COPD), Diabetes, Hypertension and Age) using transcriptomic data sets publicly available^10,28,29^. All the genes available in each dataset were first sorted under each condition, based on the log fold change (Condition vs. Control) with the top 25% genes classified as High (H), having the largest fold change (higher in Condition), and the bottom 25% genes classified as Low (L), having the smallest fold change (much higher in Control). Then, the 11 HLH genes were assessed to determine whether they are among the H or L genes in each condition.

We hypothesized that HLH genes, that may play an active role in thrombotic complications of COVID-19, are more likely to be regulated in co-morbid conditions. *RAB27A* expression was found altered in all studied conditions, while *AP3B1* expression was also found altered in all conditions except one, lung cancer (**Fig 3**). The protein encoded by the *RAB27A* gene has emerged as a central regulator of the neutrophil response through its regulation of vesicular trafficking and degranulation including azurophilic granule exocytosis^30,31^. *RAB27A* expression was low in females, aged individuals and hypertension whereas it was highly expressed in smoking, cancer and diabetes. The protein encoded by the *AP3B1*, part of the AP-3 complex that is ubiquitously expressed, mediates proteins sorting from the endosomal and trans-Golgi network to lysosomes and endosome-lysosome–related organelles^32^. *AP3B1* expression was low in females, COPD and diabetes but high in smoking, hypertension and aged individuals. It is always hazardous to infer changes in activity based on changes in gene expression, but the pattern of results shows that genes causing primary HLH have their expression regulated in many of the co-morbid conditions of COVID-19. Moreover, by looking at the clustering pattern emerging from the HLH genes, the following conditions were clustered: smoking and lung cancer, COPD and diabetes, hypertension and aging. Overall, this defines potentially shared risk for disease severity within these pairs in regard to their pathophysiology. Treatment approaches aimed at decreasing the risk of thrombosis may differ between these groups.

**Figure 3.**
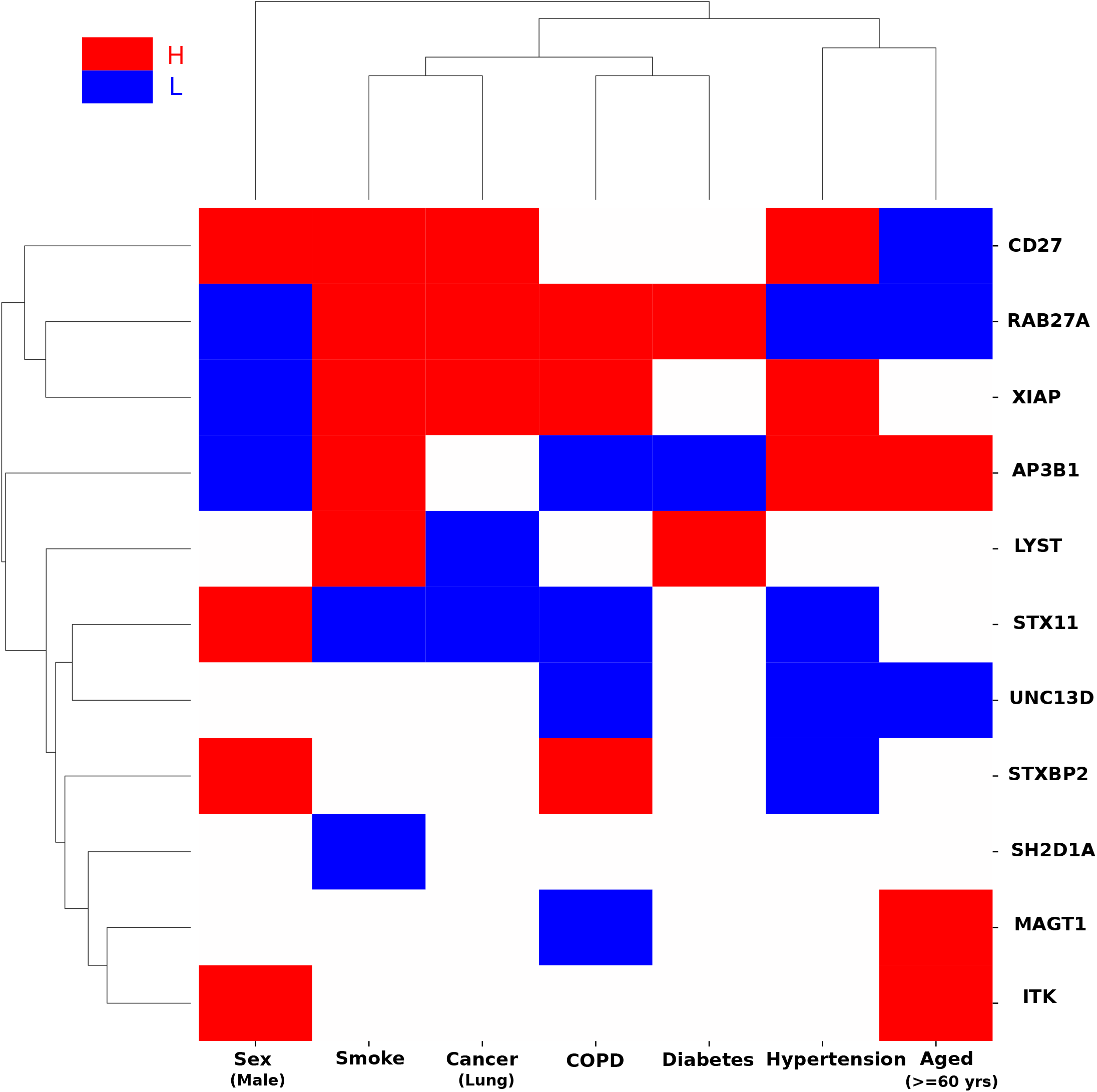
Differential expression of HLH genes in COVID19 associated health conditions. Gene expression of lung cells under different health conditions that have been associated with COVID19^10,28,29^ was analyzed. The health conditions include Sex (Male Vs. Female healthy individuals), Smoking status (current smokers vs never smokers), Cancer (Lung cancer vs. Normal), COPD (COPD vs. Normal), Diabetes (Type 1 Diabetes vs. Normal), Hypertension (Severe PH: mPAP>40mmHg vs. without PH: mPAP<20mmHg), and Aged (>=60 years vs. <60 years healthy individuals). All the genes in these datasets were sorted under each condition, based on the log fold change (condition vs control). Genes whose fold change was among the top 25% were classified as high (H) and those whose fold change was among the bottom 25% were classified as low (L). Finally, the 11 HLH genes were assessed to determine whether they are among the H or L genes in each condition. The red blocks represent HLH genes that were highly expressed (H, top 25%) in condition (vs. control) while the blue blocks represent HLH genes that were lowly expressed (L, bottom 25%) in condition (vs. control). We compared the HLH genes and all background genes in terms of H/L expression under various conditions. We first counted the number of H/L (differentially expressed between the condition and control) for each of the HLH genes, and then for each of the background genes. Next, we used a one-sided Mann-Whitney rank test to determine whether the HLH genes have larger absolute fold changes, (*i*.*e*, are differentially expressed), in COVID19 associated conditions compared to all the background genes significantly (p-value<0.05). The average number of H/L conditions for HLH genes (red or blue blocks) is 3.82, which is significantly larger (one-sided rank-sum test p-value=1.01e-4) than the average number of H/L conditions for all genes (1.70).

### HLH genes are functionally associated with neutrophils degranulation

The next crucial question was to determine what function regulated by HLH genes may be linked to the risk of thrombosis in COVID-19. Gene ontology (GO) enrichment analysis was done to identify plausible biological functions regulated by HLH genes using the KnowEnG analytical platform (https://knoweng.org/analyze)^33^. KnowEnG is a computational system for analysis of ‘omic’ datasets in light of prior knowledge in the form of various biological networks. After correcting for multiple hypothesis (Benjamini-Hochberg method), the standard mode (no network used) did not find any significant GO terms (False discovery rate (FDR)>0.05). However, the knowledge-guided mode that used three different networks (protein-protein interaction (PPI), co-expression network from the STRING database^34^ and HumanNet Integrated network^35^ identified several GO terms (**Table 1 and Supplementary Table S2**). Neutrophil degranulation was the top GO term identified with all three networks above.

**Table 1.** GO terms associated with the HLH gene set, identified by KnowEnG’s network-guided gene set characterization pipeline using HumanNet Integrated network.

| GO ID <sup>a</sup> | GO Term Description <sup>a</sup> | Difference Score <sup>b</sup> |
| --- | --- | --- |
| GO:0043312 | Neutrophil degranulation | 1 |
| GO:0006886 | Intracellular protein transport | 0.689 |
| GO:0070382 | Exocytic vesicle | 0.675 |
| GO:0045921 | Positive regulation of exocytosis | 0.672 |
| GO:1903307 | Positive regulation of regulated secretory pathway | 0.660 |
| GO:0043304 | Regulation of mast cell degranulation | 0.645 |
| GO:0032438 | Melanosome organization | 0.634 |
| GO:0006968 | Cellular defense response | 0.625 |
| GO:0031201 | SNARE complex | 0.616 |
| GO:0005764 | Lysosome | 0.613 |
| GO:0051607 | Defense response to virus | 0.611 |
| GO:0042127 | Regulation of cell proliferation | 0.610 |
| GO:0033093 | Weibel-Palade body | 0.578 |

<sup>a</sup> From KnowEnG<sup>33</sup> (www.knowng.org/analyze)
<sup>b</sup> See methods section.

While HLH is traditionally associated with hyper activation of T-cells and macrophages, an investigation published in 2019 showed that in addition to T-cells, targeting neutrophils provided benefits in a murine model, indicating an underappreciated role of neutrophils in this syndrome^36^. Other GO terms point to the importance of vesicular trafficking/exocytosis (intracellular protein transport, exocytic vesicle, positive regulation of exocytosis, positive regulation of regulated secretory pathway, regulation of mast cell degranulation, melanosome organization, SNARE complex, lysosome and Weibel-Palade body) and host defense response (cellular defense response and defense response to virus) as would be expected for genes associated with diseases that presents with coagulation and inflammation related processes. A main finding of this study is a proposed role for neutrophil degranulation in HLH and by extension in COVID-19 coagulopathies. Amongst the important functions associated with neutrophil degranulation is the release of NETs^37^.

### NET-release, a clue to the risk of SARS-CoV-2 infection complications in rheumatologic pediatric diseases?

Emerging evidence has suggested an association between KD-like illness (KD) and severe complications of COVID-19 in children^21,38^. Of note, Neutrophil Extracellular Traps (NETs) formation is increased in KD-derived neutrophils^39^. NETs are protein-coated DNA extruded from neutrophils that directly induce dose-dependent platelet aggregation and dense granule secretion^40^, playing an important role in coagulopathies ^41^. NET formation can occur via two mechanisms, either a cell death-mediated process (NETosis) occurring over a period of hours or a non-lytic release mechanism via degranulation and expulsion of chromatin that is initiated within minutes of activation^37^. HLH has been linked to NET induction and production of cytokines such as IL-1β, IL-6, IL-8 and IL-17^42^, elevated in both KD and COVID-19^43^. Reactive HLH presents clinically as primary HLH (of genetic origin) and has been described in other pediatric rheumatologic diseases where NETs could also play key pathogenic roles such as in systemic juvenile idiopathic arthritis (sJIA) and lupus^44,45^. NETs have been proposed to be part of the pathogenesis of sJIA or MAS secondary to sJIA^46^. We did not have access to transcriptomic data form KD-derived neutrophils. However, we were able to access such a dataset from sJIA^47^, to investigate the expression of the HLH-SARS-CoV-2 interactome (**Supplementary Table S1**). The majority of the genes were expressed in neutrophils and 7 of them (UNC13D, LYST, AP3B1, MAGT1, RAB8A, GOLGA2 and G3BP1) were significantly elevated in either inactive or active sJIA compared to control neutrophils (**Fig. 4**). It is worth noting that the most connected gene to SARS-CoV-2, AP3B1 that can directly interact with SARS-CoV-2 E protein, is in the list of up-regulated genes in neutrophils derived from sJIA. An important message stemming from our discovery that NETs may be drivers of coagulopathies in reactive HLH is the potential susceptibility of a subset of the pediatric population, identifying them as at risk of severe complication of COVID-19. This led us to the next step, predicting potential vulnerable populations to thrombolytics complications of COVID-19 based on their susceptibility to release NETs.

**Figure 4.**
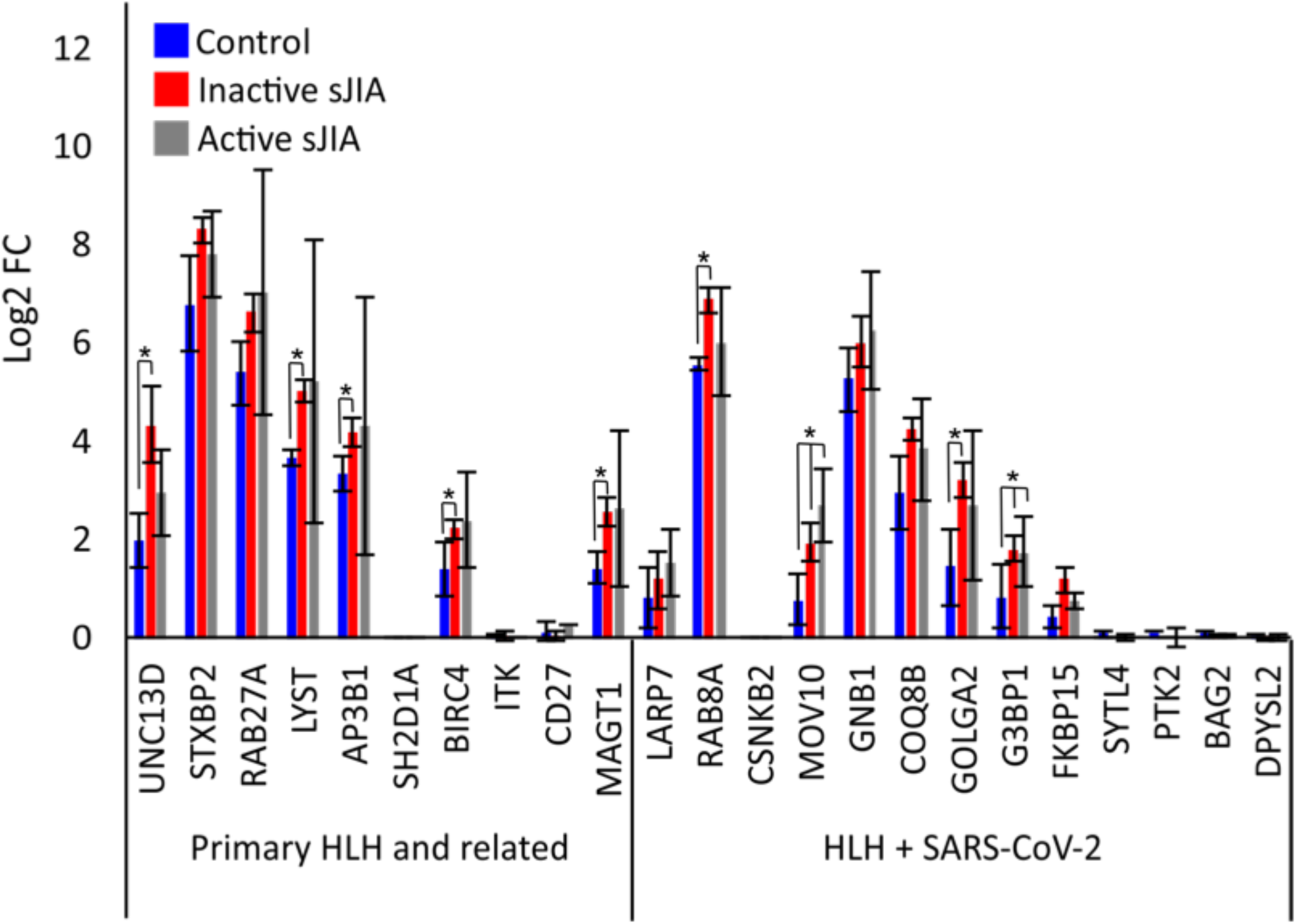
Expression of HLH genes in Control, inactive sJIA and active sJIA Neutrophils. Gene expression of HLH-SARS-CoV-2 and positive COVID-19 genes (**Supplementary Table 1**) in sJIA was calculated from GEO series GSE122552^47^. Data was mapped to the hg38 genome and normalized by reads per kilobase per million (RPKM). Values for HLH genes were displayed for control and sJIA patients that were either in remission (inactive sJIA) or had active symptoms (active sJIA).

### Identifying potentially vulnerable populations to COVID-19 based on NETs release

The World Health Organization has established that identifying vulnerable populations is an urgent public health priority in the context of the COVID-19 pandemic^48^. Based on the analyses above, NETs may play an important role in promoting thrombosis in COVID-19. The role of neutrophils in coagulopathies is becoming increasingly recognised and particularly that of NETs^41^. Therefore, we hypothesized that health conditions associated with increased release of NETs could be a predictive factor for thrombotic complications of COVID-19. Based on this hypothesis and in order to identify vulnerable populations, we developed a method called foRWaRD (in<u>fo</u>rmative <u>R</u>andom <u>Wa</u>lk for <u>R</u>anking <u>D</u>iseases) to rank different diseases associated with NETs based on their relevance to a gene set of interest (here genes in the HLH-SARS-CoV-2 network) (see Methods for details).

We obtained two NET gene signatures from previous studies^49,50^ and combined them to obtain a list of NET-associated genes (**Supplementary Table S1**). Then, we obtained the list of diseases associated with these genes from the DisGeNET database^51^. Entries with gene-disease association (GDA) <0.4, were filtered out to arrive at a set of 99 diseases. We used the set of 24 genes in the HLH-SARS-CoV-2 interaction network (**Fig. 1**) as the query set and used the HumanNet Integrated network^35^ as the gene interaction network in foRWaRD. The full list of 99 diseases, ranked using foRWaRD, is provided in **Supplementary Table S3. Table 2** illustrates the diseases with a Normalized Disease Score (NDS) greater than 0.5, meaning that these diseases are enriched above the background probabilities. Most of the 10 top-ranked diseases associated with genes linked to NETs can be sub-grouped in 4 major categories 1-immune/infectious (Alzheimer’s disease, Immunodeficiency 8); 2-cardiovascular (Myocardial reperfusion injury, Hemolytic anemia due to G6PI, Bleeding disorder type; 15); 3-metabolic (Diabetes) and 4) Cancer (Liver carcinoma). It is important to note that some of these diseases would be putatively associated with NET-deficiency such as Immunodeficiency 8, more often presenting clinically with bleeding. Whether patients suffering from these diseases are protected from thrombotic-complications of COVID-19 remains to be determined. However, diseases associated with increased NET release are expected to yield greater risk of thrombosis and may identify vulnerable populations to severe thrombotic complications of COVID-19.

**Table 2.** Diseases associated with NETs and HLH genes, identified using foRWaRD.

| Rank | Disease | Disease Score | NET<sup>a</sup> | Remarks |
| --- | --- | --- | --- | --- |
| 1 | Alzheimer's Disease | 1 | + | Increased NET in the proximity of Amyloid beta deposit<sup>83</sup>. Increase risk of thrombosis<sup>84</sup> and stroke<sup>85</sup>. |
| 2 | Liver carcinoma | 0.599 | + | NETs have been described in multiple malignancies<sup>83,55</sup>, including liver carcinoma<sup>86</sup>, which is also associated with an increased risk of VTE<sup>87</sup> |
| 3 | Myocardial Reperfusion Injury | 0.516 | + | Neutrophils play a major role in reperfusion injury that result in microvascular damage<sup>88</sup>. Reperfusion injury leads to the release of NETs in animal models<sup>89</sup>. |
| 4 | Diabetes Mellitus | 0.508 | + | In COVID-19, well-controlled blood glucose was associated with lower mortality compared to individuals with poorly controlled blood glucose<sup>63</sup>. Accordingly, proteomic analysis of peripheral blood polymorphonuclear cells (PBMCs) reveals alteration of NET components in uncontrolled diabetes<sup>90</sup>. Interestingly, high glucose cooperates with IL-6 and homocysteine to form NETs in the context of Type 2 diabetes (T2D)<sup>91,92</sup>. T2D is a major COVID-19 co-morbidity<sup>63</sup>. |
| 5 | Congenital Fiber Type Disproportion | 0.506 | (-) | None |
| 6 | Immunodeficiency 8 | 0.502 | - | Results from the loss of function of coronin-1, leads to bleeding. Coronin-1 plays key functions in PMN trafficking in innate immunity<sup>74</sup>. |
| 7 | Diabetes Mellitus, Insulin-Dependent | 0.501 | + | See entry above |
| 8 | Cataract 30 | 0.501 | (-) | None |
| 9 | Hemolytic anemia, nonspherocytic, due to glucose phosphate<br>isomerase deficiency | 0.500 | (-) | None |
| 10 | Bleeding disorder,<br>platelet-type, 15 | 0.500 | (-) | None |

Abbreviations used: NETs = Neutrophil Extracellular Traps.
<sup>a</sup> A “+” sign indicates demonstrated NET release in the disease, a “-” sign a deficiency in NETs. Brackets “( )” around the “+” or “-” signs indicate prediction without published data.

## Discussion

Based on recent literature, we hypothesized that severe pulmonary thrombotic complications of COVID-19 are associated with a hematologic cytokine storm that could be, in part, defined using genes causing HLH. The network-informed analysis presented in this paper, revealed that 1) the top GO biological function associated with HLH genes is neutrophil degranulation, consistent with a recent report highlighting the undervalued role of neutrophils in HLH^36^; 2) HLH genes are significantly enriched with the SARS-CoV-2 human interactome; 3) the top-ranked HLH gene, *AP3B1*, has roles in cargo loading of type II pneumocytes, where it may interact with SARS-CoV-2 to disturb surfactant physiological functions to promote inflammation/pro-coagulation activities; 4) diseases/syndromes-associated with increased release of Neutrophil Extracellular Traps (NETs) may predict vulnerable populations, including those affecting children.

Taken together, the network-informed analysis led us to propose the following model: the release of NETs in response to inflammatory signals acting in concert with SARS-CoV-2 damage the endothelium and direct platelet-activation promoting abnormal coagulation leading to serious complications of COVID-19 in susceptible individuals (**Fig. 5**). The underlying hypothesis is that genetic and/or environmental conditions that favor the release of NETs may predispose individuals to thrombotic complications of COVID-19 due to an increase risk of abnormal coagulation. This would be a common pathogenic mechanism amongst numerous conditions including autoimmune/infectious diseases, hematologic and metabolic disorders.

**Figure 5.**
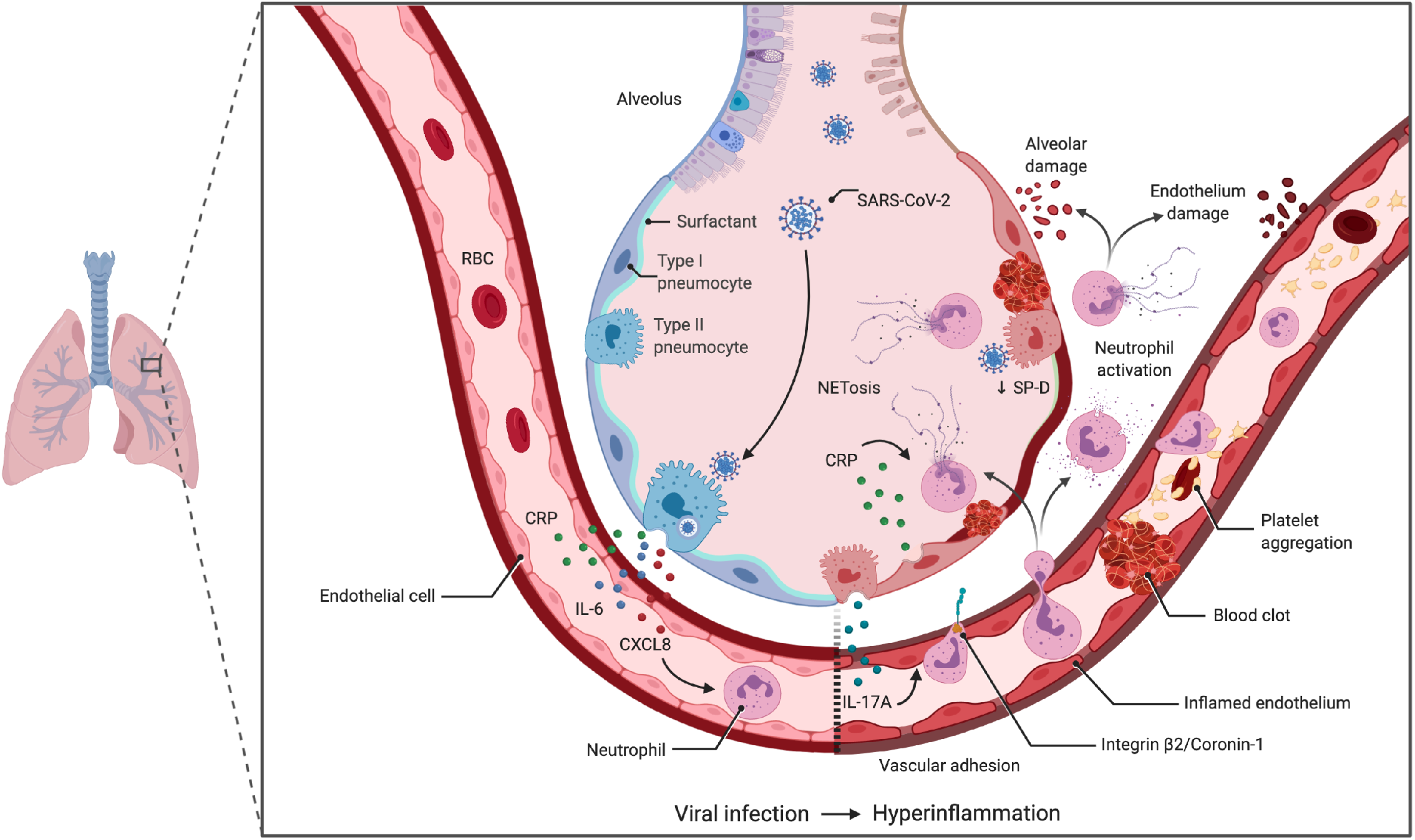
Model of NET-mediated endothelial damage contributing to pulmonary vascular thrombosis in severe COVID-19. Infection by SARS-CoV-2 in vulnerable population will lead to hyperinflammation either from underlying genetic mutations, specific epigenetic landscapes or external factors, that will result in the increase circulation of acute phase reactants such as CRP and pro-inflammatory cytokines associated with neutrophilia like IL-6, IL-17A/F and CXCL8 (IL-8). IL-17A activates the endothelium to induce neutrophil adhesion^93^, where the increase in CRP can trigger the release of NETs, resulting in damage to the endothelium as well as aggregation and activation of platelets. Additionally, the presence of SARS-CoV-2 E protein in type II pneumocytes could disturb the surfactant cargo via its interaction with AP3B1, leading to impaired secretion of SP-D and greater NET formation by septal and intra-alveolar neutrophils increasing the risk of thrombosis in the pulmonary microvasculature. In some predisposed patients the combinations of these mechanisms will lead to severe COVID-19 complications. The identification of mediators of this pro-coagulation cascade is essential in achieving the two-fold task of identifying vulnerable populations and developing a personalized medicine approach.

The role of neutrophils in coagulopathies is becoming increasingly recognised and particularly that of NETs^41^. Interestingly, elevated Neutrophils count is the best single leukocyte predictor of cardiovascular risk^52^, bettered only by the combination of high neutrophils to low lymphocytes ratio^52^, a clinical feature of COVID-19^10^. NET release can be triggered by various inflammatory mediators found elevated in severe COVID-19, including CRP, IL-1β, IL-6 and IL-8^43^. There is also a positive correlation between circulating serum of IL-6, IL-8, CRP and NET levels^53^. NETs are found in a variety of conditions such as infection, malignancy, atherosclerosis, and autoimmune diseases with reports now emerging that describe their presence in COVID-19^9,54–56^. Amongst the known diseases associated with NETs, several are related to children including Cystic Fibrosis^57^, Meningococcal Sepsis^58^; Lyme Neurobiellosis^59^; Juvenile Dermatomyositis^60^ and pediatric inflammatory bowel diseases^61^. In pediatric sepsis, NETs levels were elevated and correlated with disease severity, mirroring results in mice where higher NETs levels in response to lipopolysaccharides are found in infant mice compared to adults^62^.

One of the quickest ways to decrease the burden of COVID-19 on the health care systems throughout the world is to identify at-risk populations to emphasize the importance of infection prevention measures for those individuals. Since these measures incur high personal, social and economic costs, a precise knowledge is essential. We presented a novel computational algorithm that enabled us to identify potential diseases linked with NETs (**Table 2**). Interestingly, amongst the identified diseases, Diabetes, a well-established comorbidity of COVID-19^63^, is ranked 4^th^ and 7^th^. Our study provides additional insight into the potential mechanisms involved, with increase NETs formation resulting from the underlying chronic inflammation as a key factor promoting coagulopathies in diabetics suffering from COVID-19. As for the top ranked disease, Alzheimer’s Disease, whether NETs in the brain can lead to an increased risk of systemic thrombosis looks less likely than the reverse, that SARS-CoV-2 infection may increase NETs release in the brain that could exacerbate Alzheimer’s disease-driven pathology including a greater risk of stroke. This may be an important question for future studies due to the susceptibility and severity of the elderly to COVID-19 and notably the extreme mortality seen in long-term care home arounds the world where cognitive impairment is highly prevalent^64,65^.

It has been suggested that COVID-19 should be added to this list of hyperferritinemic syndromes, which includes adult-onset Still’s disease, septic shock, catastrophic anti-phospholipid syndrome, and MAS (reactive HLH)^66^. Collectively, these diseases may share similar underlying factors of complications, including an underappreciated role of NETs leading to coagulopathies. It is possible that individuals can unfortunately contract SARS-CoV-2 infection in addition to other factors that underlie any of these conditions (other viruses for example), which may lead to further amplification loop. PCR-negative SARS-CoV-2 patients presenting with clinical symptoms of hyperferritinemic syndrome should be considered highly vulnerable and appropriate infection control measures should be put in place.

Disorders associated with bleeding should decrease the risk of thrombotic complications of COVID-19 (however they may still lead to severe COVID-19 via other mechanism). Nevertheless, they can be informative on pathophysiology. The strongest connectivity to SARS-CoV-2 E protein was AP3B1 (**Fig. 1**). Loss of function of *AP3B1* leads to Hermansky–Pudlak syndrome type 2 that is associated with bleeding and coagulation defects^67,68^. The SARS-CoV (2002-2003 strain) E (envelop) protein is mainly localised in cells at the ER-Golgi compartment of cells^69^, where it participates in assembly, budding, and intracellular trafficking of viruses^70^. Therefore, both proteins have a coherent subcellular localization supporting their potential interaction. Moreover, in post-mortem immunohistochemical analysis of lung tissue, the SARS-CoV-2 S (spike) and E proteins were found to localize with the respiratory epithelia, the interalveolar, and the septal capillaries^5^. In addition, septal and intra-alveolar neutrophilia was observed^5^, colocalizing some of the key players of a neutrophil-driven SARS-CoV-2 enhanced coagulation cascade in COVID-19 (**Fig. 5**). Whether SARS-CoV-2 E protein can directly or indirectly penetrate neutrophils and/or platelets remains unknown, as these cells are not reported to highly express ACE2+/TMPRSS2+, the two key host proteins for viral entry. However, both of these proteins are highly expressed on type II pneumocytes^71^, where AP3B1 is important for cargo loading of lamellar bodies^72^. A postmortem examination in a COVID-19 patient who succumbed to a sudden cardiovascular accident revealed SARS-CoV-2-viruses present in pneumocytes despite PCR-negative nasal swabs^73^, indicating a prolonged risk in the lower airways for complications.

Immunodeficiency 8, resulting from the loss of function of coronin-1, also leads to bleeding. Coronin-1 plays key functions in PMN trafficking^74^ in part via its interaction with the integrin β2. β2 integrin-mediated systemic NET release is a viral mechanism of immunopathology in hantavirus-associated disease such as kidney and lung damage^75^, similar to the immunopathology in severe COVID-19. Overall, diseases associated with a putative loss-of-function of NETs suggest mechanistic roles for AP3B1, coronin-1 and integrin β2 in regulating NET-mediated coagulopathies in the lung alveolar and peri-alveolar areas (**Fig. 5**).

The analysis in this study is based on a new algorithm that we develop (freely available at https://github.com/phoenixding/genelist2covid19). GeneList2COVID19 can systematically evaluate the connection of any given gene list to SARS-CoV-2 proteins both within-host proteins and between host-viral proteins. Therefore, it can be used to study a wide variety of biological problems associated with COVID-19, especially in circumstances where experimental data on COVID-19 (e.g. transcriptomics or genomics) is not yet available for the problems of interest. The algorithm was found effective, on positive (proven to be associated with COVID-19) and negative (irrelevant to COVID-19) gene lists. In terms of limitation, GeneList2COVID19 is dependent on the prior knowledge of the protein interactome within the host, and between the host and virus. Currently, we have a well-established protein-protein interactome for the human species. However, the interactome between the SARS-CoV-2 proteins and human proteins is relatively limited^23^, since such an interactome is far from complete. For example, there are no reported interactions for ACE2 and TMPRSS2, which are critical to SARS-CoV-2 infection. We provided an option (-v) in GeneList2COVID19 to utilize any new host-viral protein interactome data when it becomes available. While, GeneList2COVID19 is good at telling whether an input gene list is associated with COVID-19, it cannot test the mechanistic hypothesis generated. It can provide the network that connects the genes in the list of the SARS-COV-2 proteins, but it cannot determine which nodes/edges in the network is more critical (and when they are activated). At this stage, the most useful information is derived from considering the entire network. As the availability of COVID-19 related “-omics” data increases, we will extend the method into a joint-model that integrates all those omics data for a more comprehensive, high-definition network model that can provide additional and more precise insights for the role of genes in COVID-19. The second computational algorithm provided, foRWaRD (https://github.com/ddhostallero/foRWaRD), is also limited by the requirement of known gene-disease associations. As a result, this method does not work with syndromes/conditions that do not have a well-established cause (and associated genes) or are from infectious origins, an important trigger of NETs formation.

## Conclusion: NETs as a determinant of severe thrombotic complications of COVID-19

The results presented herein points to neutrophils and the release of NETs as important mediators of coagulopathies in COVID-19. In a murine model of the 2002-2003 SARS-CoV infection, respiratory failure was associated with lung neutrophilia^76^. Accordingly, in a study investigating autopsy-derived lung tissue of COVID-19, septal and intra-alveolar neutrophilia was observed^5^, which could contribute to pulmonary embolism atypical from other ARDS. In addition, the levels of NETs correlate with poor prognosis in severe influenza A viral infections^77^. Taken together, this supports a central role of NETs in the risk of thrombotic complications of COVID-19, that may be particularly relevant for children suffering from underlying rheumatologic or infectious conditions. Further studies in well-defined cohorts of COVID-19 patients are mandatory to confirm the relevance of the observations highlighted in the present study. Such knowledge may be of importance in novel COVID-19 severity biomarkers identification that will be needed in the management of individuals at risk of complications.

## Methods

### Datasets

For this study we used the following datasets: The interactions between the SARS-CoV-2 proteins and host proteins^23^, reporting 332 interactions that involve 26 SARS-CoV-2 proteins and 332 human host proteins. Each interaction in the map was assigned an interaction score that represents the strength of the interaction (a score between 0 and 1). We also collected the protein-protein interactions (with interaction scores) between all human host proteins from the HIPPIE database^78^ We obtained a list of Highly/Lowly (H/L) expressed genes under different health conditions that potentially associated with COVID19^10,28,29^. To identify the vulnerable populations using foRWaRD, the full list of genes associated with these diseases were downloaded from the DisGeNET database (www.disgenet.org)^51^. We downloaded the HumanNet Integrated network^35^ from KnowEnG’s Knowledge Network version 17.06 (https://github.com/KnowEnG/KN_Fetcher/blob/master/Contents.md) and used as input to foRWaRD.

### Network-guided gene set enrichment analysis using KnowEnG

We used the gene set characterization pipeline of KnowEnG^33^ (www.knowng.org/analyze) in the standard mode and in the knowledge-guided mode to identify GO terms associated with the HLH genes. The standard mode performs Fisher’s exact test, while its knowledge-guided mode is an implementation of DRaWR^79^, a method that utilizes RWRs in order to incorporate gene-level biological networks in the enrichment analysis to improve identification of important pathways and GO terms. For the knowledge-guided mode, we used three biological networks to augment the GO enrichment analysis: the experimentally verified protein-protein interaction (PPI) and the co-expression networks from the STRING database^34^ as well as the HumanNet Integrated network ^35^. For the analysis, we did not use the bootstrapping option, selected homo sapiens as ‘species’, and used default values for all other parameters. We obtained GO terms with a “difference score” above 0.5. This score represents the normalized difference between the query probabilities and the baseline probabilities in the RWR algorithm, with the best score observed as 1 (**Table 1**).

### Building the HLH-SARS-CoV-2 interaction network

We first built a network that connects all the SARS-CoV-2 proteins and the human host proteins based on the collected protein interaction data. The edges connecting different proteins are weighted based on the interaction scores obtained from the original datasets above. Next, we inferred the signaling paths from SARS-CoV-2 proteins down to a list of proteins (genes) of interest.

A few key assumptions must be made before we can make such inference. First, since collected protein interactions within the host (and between the host and the SARS-CoV-2 virus) do not have directions, the reconstructed network graph is undirected. Here, we assumed that the information (i.e., infection) flows from the SARS-CoV-2 proteins to the proteins that directly interact with SARS-CoV-2 proteins, next to other intermediate signaling proteins, and finally to the target genes (proteins) of interest. There might be multiple intermediate proteins residing between the direct SARS-CoV-2 interacting proteins and the target proteins of interest. Second, we did not allow loops in our path from SARS-CoV-2 proteins to the target proteins to reduce the computation complexity. Although feedback loops have been reported in previous studies^80^, they are still relatively rarely observed in the human protein-protein network^81^. Last, we assumed that the interaction score between two proteins is proportional to the strength (or the likelihood) of their interaction. A larger interaction score represents either a stronger or more likely interaction, which results in a “stronger” connection edge in both cases.

The objective of the analysis was to find the strongest (or most likely) “connecting” path from SARS-CoV-2 proteins to the target proteins (genes) of interest in the constructed network, where the connection strength was quantified by a “connectivity score”. We formulated the above problem as:

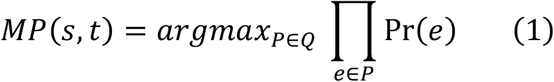

where *MP(s,t)* is the most likely path from source node *s* to the target node *t, 𝒬* is the union set of all possible paths *P* from *s* to *t*, and *e* is the edge on the path *P. Pr(e)* is the probability of the edge *e*, which is the interaction score (range [0,1]) between two proteins (ends) of the edge. Since the protein-protein interaction scores were all in the range of [0,1], we directly used them to represent the interacting probabilities. The above equation can be re-written as the following maximization problem:

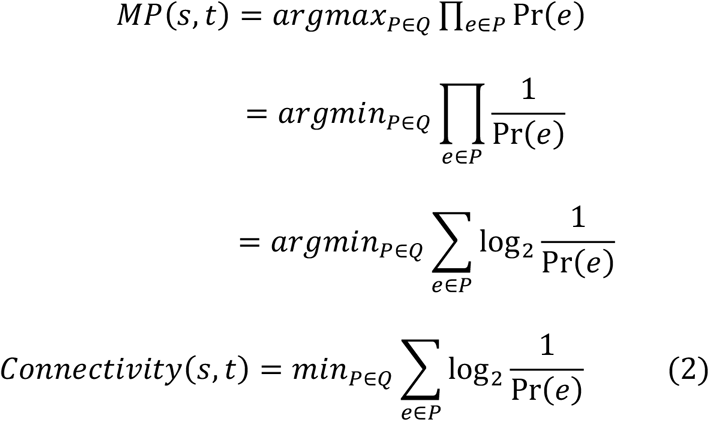

Note that *Pr (e) >0*, as all the reported interaction scores are strictly positive. This minimization problem denotes the “shortest path problem”, which we solved using Dijkstra’s algorithm (with a quadratic time complexity in the number of vertices).

The above optimization strategy relies heavily on the strong edges (interactions with high scores). The preference of high score edges may lead to over-sized paths, composed of only high score edges. To avoid oversized paths, we penalized/constrained the length of the path (# of edges in the path) while minimizing the connectivity score (a smaller connectivity score represents stronger connectivity). Here, we revised the aforementioned optimization problem into a 2-pass strategy. In the first pass, we find all the shortest paths *X(s,t)* (with the same path length) that connect SARS-CoV-2 proteins to the target proteins of interest, without considering the edge weights (interaction scores) in the graph. In the second pass, we find the path *x(s,t)* in *X(s,t)* that produces the minimal connectivity score Connectivity_constrained_ (s,t) by taking the weight scores into considerations for only the selected candidate paths from the first run.

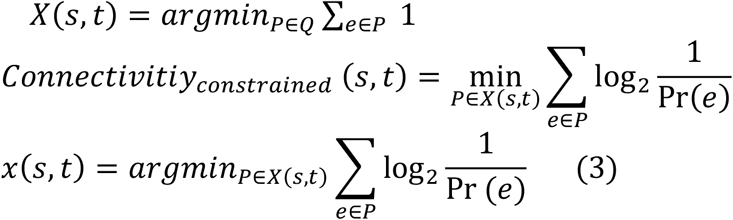

We have packaged all the code into a tool named GeneList2COVID19, which is freely available for academic uses.

### Ranking diseases using foRWaRD

We developed foRWaRD to rank a set of diseases (with known associated genes) based on their relevance to a set of genes (here HLH genes). This method works on the principles of Random Walk with Restarts (RWRs) for ranking genes and gene sets on heterogenous networks^33,79,82^, and enables integration of gene-level interactions to rank a set of diseases, with known associated genes, based on their relevance.

foRWaRD, requires three types of inputs (**Sup. Fig. 1**): 1) a set of diseases along with genes associated with each disease and the score of gene-disease associations (optional), 2) a gene interaction network (e.g. co-expression, protein-protein interaction, etc.), and 3) a set of query genes. Using these inputs, foRWaRD first generates a heterogeneous network comprising of gene-gene edges and disease-gene edges, with normalized edge weights representing the strength of the gene-gene interaction and the strength of evidence for gene-disease interaction (e.g. from the DisGeNET database). Then, the query set is superimposed on this network and is used as the restart set in an RWR algorithm. Using RWR in this algorithm allows us to capture topological information within the network both locally (the neighborhood surrounding the query set) and globally. After the convergence of the RWR, the steady-state probabilities of the disease nodes represent their relevance to the query set. In order to correct for the network bias (i.e. to avoid diseases with a large number of associated genes be ranked highly independent of their relevance to the query set), we run the RWR one more time with all the genes in the network as the restart set, providing a background steady-state probability for each disease node. The difference between the steady state probabilities of these two RWRs are then normalized between 0 and 1. More specifically, let *d*_*i*_ represents the difference between the steady state probabilities of the two RWRs for disease *i*, where *i* = 1, 2, …, *n* and *n* is the total number of diseases to be ranked (note that −1 ≤ *d*_*i*_ ≤ 1). Also, let *d*_*max*_ = *max*_*i*_ (|*d*_*i*_|). The normalized disease score (NDS) for the *i*-th disease is:

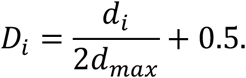

It is important to note that NDS above 0.5 reflect diseases whose similarity score with respect to the query set is larger than their similarity score with respect to all genes (i.e. background).

The RWR (which we used in foRWaRD) is an algorithm for scoring the similarity between any given node of a weighted network and a query set of nodes. Starting from some initial node, at each step the random walker moves to an adjacent node with a probability proportional to the edge weight connecting the two nodes and with some probability (known as probability of restart) it jumps to one of the nodes in the query set (also known as the restart set). The restart probability *p* controls the influence of the local topology of the network (surrounding the query set) and its global topology. We used *p* = 0.5 to balance the influence of these two factors.

### Software Availability

The software GeneList2COVID19 is written in Python, available as an open source tool at GitHub (https://github.com/phoenixding/genelist2covid19). An implementation of the software foRWaRD is available in Python and is freely available on GitHub (https://github.com/ddhostallero/foRWaRD). These GitHub repositories include the source code as well as detailed instructions on how to install and use the methods.

## Data Availability

All datasets used in this study are publicly available. Moreover, the software GeneList2COVID19 is written in Python, available as an open source tool at GitHub (https://github.com/phoenixding/genelist2covid19). An implementation of the software foRWaRD is available in Python and is freely available on GitHub (https://github.com/ddhostallero/foRWaRD). These GitHub repositories include the source code as well as detailed instructions on how to install and use the methods.

https://github.com/ddhostallero/foRWaRD

## Acknowledgements

This work was partly supported by the NSERC Discovery grant RGPIN-2019-04460 (AE), and a grant from McGill Initiative in Computational Medicine (MiCM) and McGill Interdisciplinary Initiative in Infection and Immunity (MI4) (AE and SR). SR is supported by a Chercheur-Boursier Senior salary award from the Fonds de Recherche du Québec – Santé. KT is supported by a Chercheur-Junior 1salary award from the Fonds de Recherche du Québec – Santé.

## Authors’ contributions

JD, GF, AE and SR contributed to the conception or design of the work; JD, DEH, MREK, GF, NN,MGB, AE and SR contributed to the acquisition, analysis, or interpretation of data; JD, DEH, MREK, and AE contributed to the creation of new software used in the work; JD, GF, SM, JS, ND, MS, KT, AE and SR have drafted the work or substantively revised it. All the authors have approved the submitted version have agreed both to be personally accountable for the author’s own contributions and to ensure that questions related to the accuracy or integrity of any part of the work, even ones in which the author was not personally involved, are appropriately investigated, resolved, and the resolution documented in the literature.

